# A new method for bedside determination of effective lung volume (ELV) and functional residual capacity (FRC)

**DOI:** 10.1101/2025.06.26.25330343

**Authors:** Andras Gedeon, Jakob Jansson, David Patrickson, Mats Wallin

**Affiliations:** Mincor AB Stockholm Sweden; Spirotronic AB Stockholm Sweden; eHeart AB Stockholm Sweden; Department of Physiology and Pharmacology, Karolinska Institutet, Stockholm, Sweden

## Abstract

Established methods of measuring FRC involve sophisticated equipment and elaborate procedures. Here we present a new method based on CO_2_ rebreathing that has a simple fast procedure and only requires end-tidal CO_2_ monitoring.

Ten healthy subjects with diverse anthropometric and respiratory parameters were studied in the sitting position. Reference FRC (RefFRC) and tidal volume (TV) were measured with a Cosmed Quark PFT/ DLCO unit using the single-breath methane dilution technique in combination with spirometry. Rebreathing through a dead space of precisely known volume and recording the rising end-tidal CO_2_ value of the first two breaths allows the determination of ELV and the calculation of FRC. Two sets of measurements were made on each subject 15 minutes apart.

Bland-Altman analysis of a comparison between FRC and RefFRC shows a mean bias= 0.08 l, limits of agreement (LoA) of (+1.35 to −1.19; 95%CI) l. and a percentage error of PE= 0.54. When the mean value of two observations from a subject (meanFRC) is compared to RefFRC we get a mean bias = −0.04 l (LoA) of (+0.76 to – 0.83; 95%CI) l and PE=0.23

The FRC data obtained has good absolute accuracy. A mean of two observations has sufficient precision to meet the criteria for exchangeability with reference. The simplicity of the equipment and the procedure could make this method attractive both in the pre-operative and the post-operative setting as well as in out of hospital applications.

## Introduction

Pre-operative measurement of FRC, particularly in high-risk patients, is commonly used to help guide anaesthesia but also to predict postoperative pulmonary complications [1]. Post-operative monitoring of FRC allows assessment of lung recovery and detect early pulmonary complications. Although there are several well-established methods for measuring functional residual capacity (FRC) [1–4] they all require expensive, sophisticated equipment and elaborate time-consuming procedures and none of them can therefore be used routinely at bedside. They are also mostly unavailable for primary care management of common pulmonary diseases [5]. Here we describe and evaluate a new method, employing a short period of rebreathing and end-tidal carbon dioxide monitoring, to measure the effective lung volume (ELV). ELV for carbon dioxide comprises of two compartments, the gas compartment (FRC) and the lung tissue compartment. FRC is obtained from ELV using the known relationship between these compartments [6–8].

## Methods

### Ethical Approval

This study has been approved by the Swedish Ethical Review Authority on May 6 2024. It is registered as Nr 2024-02101-01. The decision is based on judgement from a panel of legal and medical professionals. This approval assures conformity with the lastest version of the Declaration of Helsinki and with other ethical requirements including those on written informed consent and consent to publish as well as data handling and storage procedures.

#### Physical and respiratory characteristics

Ten subjects, 4 women and 6 men, with no known cardio-pulmonary disease, were recruited and studied in the sitting position. They were between 39-70 (mean 55) years old, with weights between 62-99 (mean 80) kg, tidal volume (TV) was between 0.58-1.7 (mean 1,1) l and respiratory rate (RR) between 4.6-16.8 (mean 11) breath/minute. For women average RR and TV was 8.9 breath/mn and 1,2 l and for men 9.7 breath/min and 1.1 l.

#### Description of the equipment and the procedure

The equipment used for this study is shown in Fig 1. It is made up of a nose clip, a dead space volume (DV), a sampling IR CO2 analyzer, ET600 Side stream CO2CGM Module (Ronseda Electronics Co. Ltd China) and a laptop computer for data collection. TV and the functional residual capacity reference (RefFRC) were measured in the sitting position with a Cosmed Quark PFT device with a DLCO module (COSMED SRl Italy). The single-breath methane dilution method [9] was used to measure total lung capacity (TLC) and this combined with spirometry determinations of lung volumes allowed the calculation of RefFRC.

**Fig 1.**
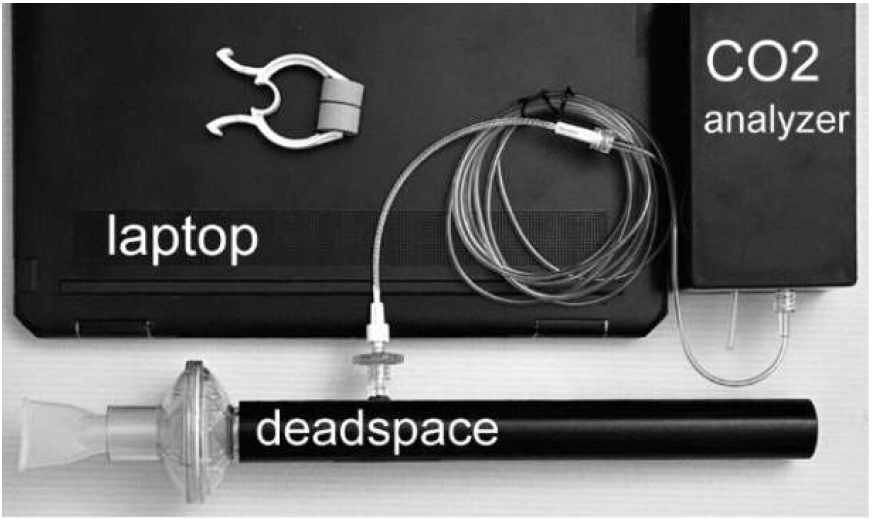

The dead space volume is formed by connecting together a mouthpiece, a respiratory filter and a pipe. The total volume is determined by an accuracy of better than 3 ml. The sampling port of the analyzer is connected to the pipe through a nafion tube and a sampling filter. DV = 0.148 l was used throughout and resulted in a DV/TV ratio in the range ~ 0.10 - 0.25. The sampling point was ~ 10 mm inside the lumen of the pipe.

The subjects were instructed to sit still for about 5 minutes. Then they were asked to apply the nose clip and to start breathing in the dead space, beginning with an expiration and to breathe in a normal manner till instructed to stop. Data collection took less than 30 seconds. After sitting still for about 15 minutes this procedure was repeated for a second set of measurements. FRC and REfFRC were measured by separate persons in separate locations and the results were blinded to each other. RefFRC was measured between the two FRC measurements.

Six subjects could be tested twice while three measurements could not be reliably evaluated due to irregular breathing and/or leakage at the mouthpiece. Thus, we obtained 17 measurements including 6 pairs of data points.

In our approach [10], first the volume of CO_2_ within the dead space is determined at the end of expiration. The CO_2_ volume in the dead space is calculated as the product of the known dead space volume and a representative sample of the CO_2_ partial pressure within the dead space at the end of expiration.

With DV/TV < 0.25 the CO_2_ partial pressure within the dead pace will decrease slowly and linearly away from the subject. At a sampling point that divides the dead space into two equal parts we can therefore get a representative average value (avPet) for the entire dead space. The CO_2_ volume is therefore obtained as a product of avPet and DV. Introducing this CO_2_ volume into ELV produces an increase in Pet, delta Pet given by

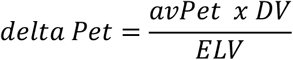

and so

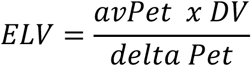

Rebreathing produces rising Pet with time as shown in Fig 2, first linearly then gradually approaching a plateau.

**Fig 2.**
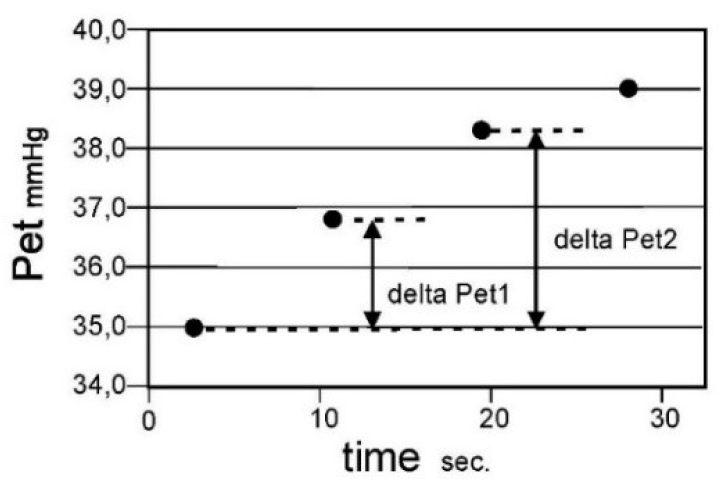

**Fig 3.**
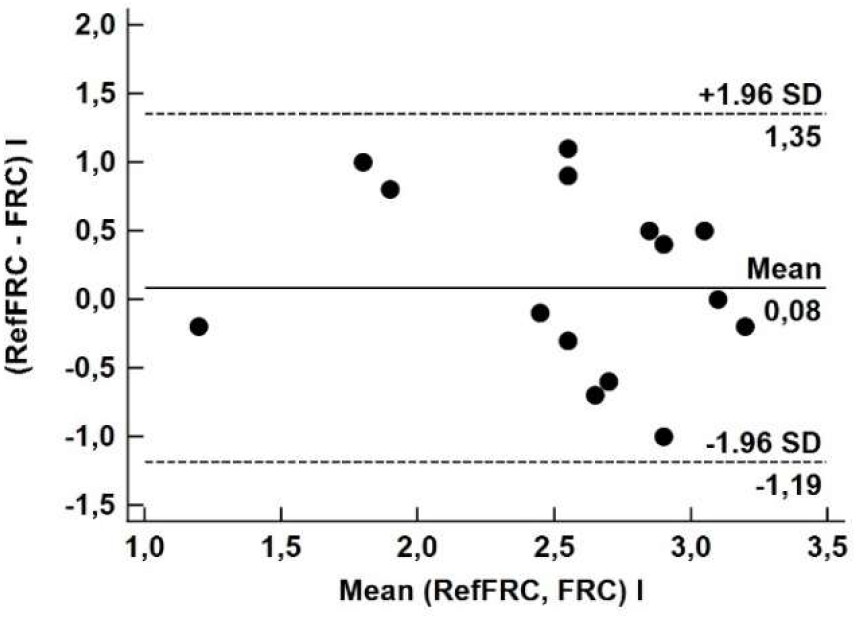

**Fig 4.**
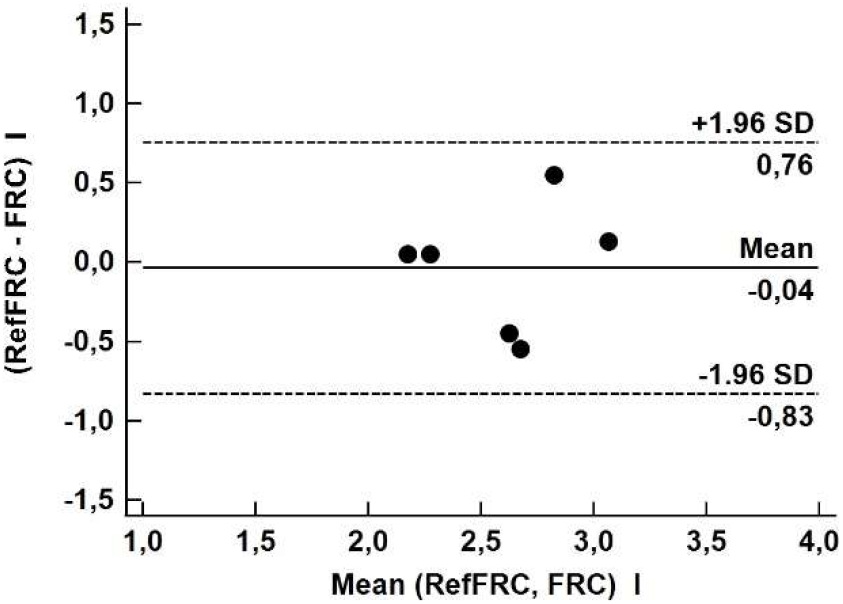

We consider only the first two breaths, which represent the nearly linear portion of the curve. In principle, the very first breath could determine the value of delta Pet but in order to get a more robust estimate that is not dependent on one data point only, we use a weighted average of the first two breaths.

The relative weight of the second breath is estimated so that it takes into account that this data point no longer falls entirely on the linear part of the curve.

We calculate delta Pet from the expression.

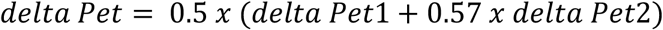

and so ELV from

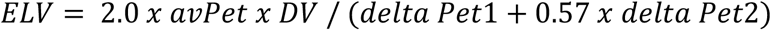

In healthy subjects, it has been shown that there is linear relationship between FRC and ELV [6] irrespective of anthropometric data. The constant of proportionality is found to be 0.80 and so we get.

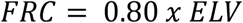

FRC obtained from this equation is compared to the reference value RefFRC.

#### Statistics

All statistical calculations were done with the MedCalc Statistical Software from MedCalc Software Ltd (Belgium) To show criterion validity for the new method we used the Bland-Altman procedure when comparing to the reference and paired t-test for comparing the two sets of measurements of FRC. Unpaired t-test was used when comparing how women and men compared to reference.

## Results

Fig 3 shows a Bland-Altman analysis [11] of a comparison between 17 observations (two identical) of FRC and Ref FRC with mean bias = 0.08 l and (LoA) of (+1.35 to −1.19; 95%CI) and percentage error PE= 0.54 Comparing the two sets of measurements with paired t-test we get a difference of 0.20 l with (LoA) of (1.09 to −0,69; 95%CI) l with t=0.58 and two-tailed probability P=0.59. Taking the difference of the women and the men data relative to RefFRC and analyzing with unpaired t-test we get a difference of −0.17 l with (LoA) of (1.52 to −1.87; 95%CI) l and t= −0.28 with P=0.79

Fig 4 shows a Bland-Altman analysis of meanFRC vs RefFRC showing a mean bias = −0.04 l (LoA) of (+0.76 to – 0.83; 95%CI) l and PE=0.23. Thus, the improved precision of meanFRC meets the exchangeability criteria of Critchley [12], PE <0.30.

## Discussion

Our method [10] is based on first principles (Dalton’s law). The only instrument used, the CO_2_ analyzer, need not be precisely calibrated because when calculating ELV the gain factor will appear both in the numerator and in the denominator and thus take each other out. ELV is determined with about the same absolute accuracy as the dead space volume, DV.

However, the accuracy of FRC, depends also on how well the constant of proportionality (K=0.80) in the equation FRC = 0.80 × ELV is known. The value we use, K=0.80 is based on a comprehensive study [6] of 90 healthy subjects and is also supported by other work [7,8]. Notably, K is found to be independent of anthropometric parameters which could point to a common, intrinsic characteristics across individuals such as for instance the volume normalized mean thickness of the lung tissue in which carbon dioxide equilibrates.

To check if K=0.80 is consistent with the known properties of an average adult lung, consider a typical case with a lung having a surface area of 90 m^2^ and a tissue volume of 600-800 ml [7]. This corresponds to a mean tissue thickness h ~ 6.7-8.9 um. If we represent FRC with a single, average sized alveoli, a sphere with radius 125 um and ELV with the same sphere but covered with a tissue layer with thickness h, and calculate the ratio between the volumes of the two spheres we get FRC/ELV = 0.79-0.84 indicating that K=0.80 is entirely consistent with the characteristics of a typical lung.

Earlier work that uses CO_2_ rebreathing to measure FRC have been oriented towards mechanically ventilated patients [13]. Contrary to our approach of using the initial rise in CO_2_ (CO_2_ wash-in), here FRC was determined from the fall of CO_2_ (CO_2_ wash-out) when returning to normal breathing subsequent to a ~ 40 s long rebreathing period. The FRC values reported, using K= 0.72, had acceptable accuracy and precision compared to both body plethysmography and nitrogen washout reference measurements. These results required stable tidal ventilation and together with the long rebreathing period made this approach mainly suitable for patients on controlled ventilation. Nevertheless, they clearly demonstrated the sound foundation of the rebreathing technique for FRC measurements.

Little is known about to what extent the equation FRC = 0.80 × ELV, valid in health, could also be used for lungs in disease. However, some indications emerge from a recent study [14] involving 45 mechanically ventilated patients all with severely reduced FRC where 25 of the patients had COVID-related ARDS. ELV was measured with the capnodynamic methodology [14] and FRC with CT-imaging. One found that non-ARDS subjects had K~ 0.88 while for patients with severe ARDS had K~ 0.77. These results suggest that the value of K is only moderately affected by disease.

We speculate that the modest difference between the two disease groups and that the K values for both groups deviate less than 10% from K=0.80, could be understood by remembering that pulmonary capillary blood has a typical volume of 70 - 90 ml while lung tissue volume is usually around 700 ml and that the amount of CO_2_ dissolved in lung tissue is about five times that contained in pulmonary capillary blood.

The precisely known dead space volume DV, has a key role in our method. DV should be neither too small nor too large compared to the tidal volume (TV) of the subject tested. If it is too small, then the increase in the Pet value will be correspondingly too small, leading to reduced precision. If it is too large, then the first two breaths used in the analysis could extend too far beyond the linear portion of the Pet curve introducing systematic errors and reducing accuracy. Too large DV can also jeopardize the assumption that the level of carbon dioxide falls off linearly within the dead space and so the sampling point dividing DV into two equal parts may no longer give a representative Pet-value for the whole DV. We have found that if DV is chosen so that the ratio DV/TV is in the range 0.1-0.25 then these pifalls are avoided.

The two sets of measurements are shown not to be statistically dependent. The subjects represented a wide range of respiratory parameters but as a group women and men were well matched. No difference in the accuracy of the method could be shown for women and men.

The major contributor to the variability in the FRC is irregular breathing. Therefore, careful attention should be given to setting up testing conditions that favor regular breathing. However, precision can be much improved by repeating the measurement and using a mean value instead of a single observation. We find that if two values are averaged, the percentage error is reduced by about a factor of two, making averaged observations exchangeable with the reference method [12].

In the present study we have chosen to repeat the measurement after 15 minutes, to ascertain that the gas exchange changes introduced by the first measurement had no influence on the second. However, since only two breaths are needed to obtain the ELV value, the gas exchange balance will be only minimally altered by a measurement and so the rebreathing maneuver could have been repeated at least as often as every minute.

## Conclusions

FRC is measured with good absolute accuracy when compared to the established reference method of single-breath methane dilution in combination with spirometry. The measurement takes only a short time (less than 30 s) and can therefore be conveniently repeated. Using the mean value of two observations increases precision so that the present method becomes exchangeable with the reference method.

The simplicity of the procedure and the equipment could make our method attractive both at bedside in the pre-operative and post-operative settings and potentially also in out of hospital environments where a measurement of FRC could add a useful tool when managing lung disease [5,15,16].

## Data Availability

All data produced in the present work are contained in the manuscript

## Notes

### Competing Interest Statement

Andras Gedeon is an employee and shareholder of Mincor AB. Mincor AB has provided economic support to this work.
No other author has any competing interest financial or otherwise

### Clinical Trial

Dnr 2025-01936-02 at the Swedish Ethical Review Authority

### Funding Statement

This study was funded by Mincor AB

### Author Declarations

This work was approved by the Swedish Ethical Review Authority. With expert groups of medical and legal professionals it covers all aspects. The study is registered at the Authority.

### Summary of Updates

Corrected name of the reference method Expanded material with Fig 4 Expanded statistics Expanded discussion section

